# Natural Language Processing Techniques to Detect Delirium in Hospitalized Patients from Clinical Notes: A Systematic Review

**DOI:** 10.1101/2025.06.05.25329038

**Authors:** Ravi Shankar, Janani Kannan, Yih Hng Tan, Xu Qian

## Abstract

**Background:** Delirium is a serious and common condition in hospitalized patients, associated with increased morbidity, mortality, and healthcare costs. Early detection and management of delirium is crucial for improving patient outcomes. Clinical notes contain valuable information about a patient’s mental status that may not be captured by structured data alone. Natural language processing (NLP) techniques have the potential to automatically detect signs and symptoms of delirium from free-text clinical notes, which could aid in early identification and prompt treatment.

**Objective:** The objective of this systematic review is to summarize and critically appraise the existing research on NLP methods for detecting delirium in hospitalized patients from clinical notes.

**Methods:** The review will follow the Preferred Reporting Items for Systematic Reviews and Meta-Analyses (PRISMA) guidelines and use the PRISMA-P framework for protocol development. A comprehensive search of PubMed, Web of Science, Embase, CINAHL, MEDLINE, The Cochrane Library, PsycINFO, and Scopus will be conducted from each database’s inception to February 2025. The search strategy will include terms related to delirium, natural language processing, machine learning, and clinical notes. Two independent reviewers will screen titles, abstracts, and full texts for eligibility using Covidence software and extract data using a standardized form. Risk of bias will be assessed using the PROBAST tool for prediction studies and the TRIPOD checklist for reporting quality. A narrative synthesis of the findings will be provided.

**Discussion:** This review aims to consolidate the current evidence on NLP approaches for delirium detection from clinical notes, identify promising methods and limitations, and highlight areas for future research and development. The findings may guide the development of automated tools to enhance the early recognition and treatment of delirium in hospitalized patients. The protocol follows best practices for systematic reviews and will be openly disseminated.

**PROSPERO Registration:** CRD42025634871

## Introduction

### Background

Delirium is an acute disorder of attention and cognition that is common among hospitalized patients, with reported incidence rates ranging from 14% to 56% depending on the clinical setting and patient population [1-3]. It is characterized by an acute onset of inattention, disorganized thinking, and altered level of consciousness that tends to fluctuate over the course of the day [4]. Delirium is associated with a wide range of adverse clinical outcomes, including increased length of stay, functional decline, institutionalization, mortality, and healthcare costs [5-8].

Despite its high prevalence and impact, delirium remains significantly underrecognized in clinical practice. A systematic review found that delirium goes undetected in 60-80% of cases in hospitalized patients [9]. Several challenges contribute to the underdiagnosis of delirium, including its fluctuating course, its overlap with dementia and depression, the lack of routine monitoring, and the underappreciation of its significance by healthcare providers [10-12].

The current standard for delirium identification is a comprehensive clinical evaluation by a trained provider, using diagnostic criteria such as the Diagnostic and Statistical Manual of Mental Disorders (DSM-5) [13] or screening tools such as the Confusion Assessment Method (CAM) [14]. However, this approach is time-consuming, resource-intensive, and relies on provider skills and judgment [15]. Furthermore, delirium assessments may not be consistently documented in the medical record, particularly for subtle or subsyndromal cases [16].

Clinical notes, such as progress notes, nursing assessments, and discharge summaries, provide a rich and untapped source of information about a patient’s cognitive status, behavior, and psychomotor symptoms over time. However, this information is typically recorded in an unstructured, free-text format that is not amenable to automated analysis using traditional methods. Natural language processing (NLP), a field of artificial intelligence focused on enabling computers to understand and manipulate human language,[17] offers a promising approach to unlock the potential of clinical notes for delirium detection.

NLP methods have been increasingly applied to a wide range of clinical applications, including identifying patient phenotypes,[18] extracting medication information,[19] classifying radiology reports,[20] and predicting clinical outcomes [21]. A growing body of research has also explored the use of NLP techniques to detect delirium and its associated features from electronic health records (EHRs) [22-25].

NLP approaches to delirium detection have ranged from simple keyword searches to more sophisticated machine learning algorithms that can automatically learn patterns and combinations of features associated with delirium from a labeled training dataset [26]. Rule-based NLP methods use pre-specified clinical cues and linguistic patterns (e.g., “confused,” “delirious,” “disoriented”) to flag cases of suspected delirium [27]. Machine learning NLP methods, such as support vector machines, decision trees, and neural networks, inductively learn to weigh and combine input features to optimize classification performance on a training set and can often achieve higher accuracy than rule-based approaches [29-31].

To date, no studies have systematically assessed the quality or risk of bias of individual studies, nor have they focused specifically on hospitalized patients or clinical notes. Given the rapid evolution of NLP methods and their application to delirium, there is a clear need for an updated, rigorous, and comprehensive systematic review.

### Rationale

Delirium is a critical issue for hospitalized patients, with major implications for both individual outcomes and health system costs. Early recognition is essential for prompt management and prevention of complications. However, current approaches to delirium detection that rely on clinical assessments and diagnostic coding alone are inadequate, missing a substantial proportion of cases.

Harnessing the wealth of unstructured clinical information locked in free-text notes using NLP represents a promising complementary approach to enhancing delirium detection. NLP-based delirium detection models could automatically scan admission notes, nursing assessments, and daily progress notes in real-time, flagging high-risk patients for further evaluation. This could help prompt earlier and more consistent screening, diagnosis and intervention for delirium.

A systematic review is needed to examine the large and expanding evidence base for NLP applications in delirium detection. Prior reviews have been narrow in scope or are now outdated given the rapid progress in the field. A comprehensive and critical synthesis of the available evidence is needed to inform future research and development efforts, identify common challenges and limitations, and lay the groundwork for translating models into practice.

### Objectives

The overall objective of this review is to systematically identify, characterize, and appraise the body of research on NLP methods for detecting delirium in hospitalized patients from clinical notes.

Specific aims include:

1. To determine what types of NLP techniques have been used for delirium detection from clinical text (e.g. rule-based, machine learning, deep learning)
2. To examine the performance of NLP methods for delirium detection in terms of standard metrics (e.g. sensitivity, specificity, F1 score)
3. To explore differences in NLP approaches and performance across healthcare settings (e.g. intensive care unit vs. general ward), patient populations (e.g. older adults, surgical patients), and clinical note types (e.g. nursing notes, discharge summaries)
4. To identify challenges, limitations, and areas for improvement in the application of NLP for delirium detection
5. To highlight promising directions for future research and development of NLP-based delirium detection systems

## Methods

This systematic review will be conducted in accordance with PRISMA guidelines [32] and reported using the specifications of the PRISMA-P protocol extension statement [33].

### Eligibility Criteria

Studies will be assessed for inclusion based on the following PICOS criteria:

- Population: Hospitalized adult patients, including general ward and intensive care settings. Studies focusing exclusively on pediatric, emergency department, or long-term care populations will be excluded.
- Intervention/Exposure: Application of NLP techniques to unstructured clinical notes (e.g., progress notes, nursing assessments, discharge summaries) for the purpose of detecting delirium. Both rule-based and machine learning approaches will be included. Studies applying NLP only to other data sources (e.g., radiology reports, medication lists) or for non-delirium indications will be excluded.
- Comparators: Standard methods for delirium identification, including expert clinical assessment, diagnostic codes, and validated screening tools (e.g., CAM, 4AT).
- Outcomes: Performance metrics for delirium detection, including sensitivity, specificity, positive predictive value, negative predictive value, F1 score, and AUROC. Secondary outcomes of interest include model features, annotation process, and error analysis.
- Study Designs: Original research studies developing or validating NLP models for delirium detection, including derivation, internal validation, and external validation studies. Comparative effectiveness studies of multiple NLP approaches will also be included. Reviews, commentaries, editorials, and non-peer reviewed sources (e.g. preprints) will be excluded.

We will not restrict by publication date or geography. Conference abstracts will be excluded unless a full paper is available.

### Information Sources and Search Strategy

We will search the following electronic databases from inception to February 2025:

- PubMed (NLM)
- Web of Science (Clarivate)
- Embase (Elsevier)
- Cumulative Index to Nursing and Allied Health Literature (CINAHL)
- Cochrane Central Register of Controlled Trials (CENTRAL)
- PsycINFO (EBSCO)
- Scopus (Elsevier)

A medical librarian will be consulted to develop the search strategy, which will include both controlled vocabulary terms (e.g., MeSH) and keywords related to delirium, natural language processing, and clinical notes. The search strategy will be first developed in PubMed and then adapted to the other databases. The searches will be updated shortly before submission of the final review draft to ensure capture of any new relevant citations.

Search string template:

((‘delirium’ OR ‘confusion’ OR ‘acute confusional state’ OR ‘acute brain syndrome’ OR ‘acute brain dysfunction’) AND (‘natural language processing’ OR ‘NLP’ OR ‘text mining’ OR ‘machine learning’ OR ‘deep learning’ OR ‘artificial intelligence’) AND (‘clinical notes’ OR ‘progress notes’ OR ‘nursing notes’ OR ‘discharge summaries’ OR ‘electronic health records’ OR ‘EHR’))

We will supplement electronic database searches with manual screening of reference lists of included studies and pertinent review articles. We will also contact field experts to identify additional studies, including ongoing or unpublished work. We will search for preprints in arXiv and bioRxiv. Grey literature searches will be conducted using Google and Google Scholar.

### Study Records

#### Data Management

Search results will be deduplicated and managed using Covidence systematic review software (<u>www.covidence.org</u>). Covidence will be used to coordinate the independent study screening and data extraction process.

#### Study Selection Process

Two reviewers will independently screen titles and abstracts of search results against the pre-specified eligibility criteria. Discrepancies will be resolved by consensus or by a third reviewer. Full texts of studies deemed potentially eligible will then be independently assessed by two reviewers, with disagreements handled as above. Reasons for exclusion of full-text articles will be recorded. A PRISMA flow diagram will illustrate the study selection process.

#### Data Collection Process

A standardized data extraction form will be developed in Covidence, piloted on 5 studies, and refined as needed. Two reviewers will independently extract data from each included study. Discrepancies will be resolved by consensus or by a third reviewer if needed. Study authors will be contacted regarding missing or unclear information.

#### Data Items

The following data items will be extracted:

- General study characteristics (e.g., authors, year, country, funding source)
- Study design (e.g., model derivation, internal validation, external validation)
- Dataset used (e.g., data source, time period, sample size, inclusion/exclusion criteria)
- Patient population (e.g., general ward, ICU, age, delirium prevalence)
- Clinical note details (e.g., note types, total notes, average notes per patient)
- Delirium definition (e.g., diagnostic criteria, screening tool, diagnostic codes)
- NLP methods (e.g., rule-based, feature engineering, traditional machine learning, neural networks)
- Model details (e.g., algorithm, software, features, pre-processing steps, hyperparameters)
- Model performance (e.g., sensitivity, specificity, PPV, NPV, AUROC, F1 score)
- Other model characteristics (e.g., most important features, error analysis)
- Implementation considerations (e.g., computational efficiency, interpretability, generalizability)

#### Outcomes and Prioritization

The primary outcome will be model performance for delirium detection, based on standard metrics such as sensitivity, specificity, AUROC, and F1 score. These metrics were chosen to allow comparisons across different modeling approaches and study populations. AUROC provides a summary measure of discrimination that is independent of the threshold chosen. F1 score is the harmonic mean of precision and recall, providing a balanced assessment of model performance.

Secondary outcomes of interest include characterization of the most informative features for delirium detection, analysis of errors and discordance with reference standards, assessment of model calibration and clinical utility, and appraisal of implementability and generalizability. We will also assess reporting quality and risk of bias of individual studies.

Given the anticipated heterogeneity in study designs, settings, and modeling approaches, we will not designate a single measure as a primary outcome that must be reported universally. However, for each performance metric we will specify a preferred calculation (e.g., cross-validation estimate rather than split-sample estimate; separate estimates in derivation and validation sets rather than combined). Where necessary, we will calculate metrics from raw confusion matrix data.

#### Risk of Bias and Reporting Quality

We will use the Prediction model Risk of Bias Assessment Tool (PROBAST)[35] to assess risk of bias in individual studies with respect to 4 domains:

1. Participants: potential for selection bias or non-representativeness of the study population
2. Predictors: potential for information bias related to predictor variables used in the NLP model
3. Outcome: potential for bias in the determination of the delirium outcome
4. Analysis: potential for bias in the data pre-processing, model development, evaluation and reporting

The instrument includes 20 signaling questions across these domains that are rated as yes, probably yes, probably no, no or no information. An overall judgment of low, high, or unclear risk of bias is then made for each domain, as well as an overall judgment for the study. Two reviewers will independently apply PROBAST to each study, with disagreements resolved by consensus or a third reviewer.

In addition, we will use the Transparent Reporting of a multivariable prediction model for Individual Prognosis or Diagnosis (TRIPOD) checklist [37] to evaluate the completeness of reporting in each study. The checklist includes 22 items that cover key elements such as study objectives, data sources, participant eligibility criteria, outcome definition, predictors, sample size, missing data, model development, model performance, and model validation.

Two reviewers will independently assess each study against the TRIPOD checklist, assigning a rating of fully reported, partially reported, not reported, or not applicable to each item. Discrepancies will be resolved by consensus or a third reviewer. Adherence to the TRIPOD reporting guidelines will be summarized descriptively and may also inform judgments of risk of bias in the analysis domain of PROBAST.

#### Data Synthesis

Given the expected diversity of NLP methodologies, study settings, and patient populations, we anticipate that formal meta-analysis of performance metrics will not be appropriate. Instead, we will conduct a narrative synthesis with tabular and graphical summaries of key characteristics and findings of included studies.

We will group studies by clinical setting (e.g., general ward vs. ICU), patient population (e.g., elderly vs. adult), note types analyzed (e.g., nursing vs. physician vs. multi-disciplinary notes), and NLP methodology (e.g., rule-based vs. feature-engineered machine learning vs. deep learning). Within each group, we will describe patterns of model performance and compare across subgroups. We will qualitatively explore potential reasons for observed heterogeneity, such as differences in delirium prevalence, note quality, or cohort characteristics.

We will present paired forest plots of sensitivity and specificity for each reported model or validation exercise, with corresponding 95% confidence intervals. Where reported, we will also present forest plots of AUROC and F1 scores. These plots will be stratified by the groupings described above.

Model features (i.e., predictors) that are identified as most important for classification across multiple studies will be highlighted. Common themes in error analysis will be summarized. Attention will be called to recent innovations and advancements in NLP modeling for delirium detection over time.

Formal narrative synthesis techniques, such as thematic analysis, concept mapping, and idea webbing, will be used to organize and interpret the extracted data. We will use the synthesis to generate new insights and research questions beyond those identified in the primary studies. Quantitative data will be integrated with qualitative findings to provide a more complete and nuanced understanding of the topic.

#### Meta-Bias(es)

We will assess for potential publication bias by comparing results of highly-prominent studies versus those in smaller journals and the grey literature. We will also compare US versus non-US studies and those with and without industry funding.

We will assess for selective reporting bias by comparing the outcomes and analyses pre-specified in study protocols or registration records (if available) versus those ultimately reported in the final study publication.

We will assess for potential bias due to extracted data imbalance via a sensitivity analysis restricted to studies that report all performance metrics of interest (sensitivity, specificity, AUROC, F1).

If there are sufficient studies, we will explore potential publication bias using funnel plots of model performance metrics against their precision. Asymmetry in the funnel plot may indicate bias related to sample size or model overfitting.

#### Confidence in Cumulative Evidence

We will use the Grading of Recommendations, Assessment, Development and Evaluation (GRADE) framework to assess the certainty of evidence for each main outcome [36]. GRADE categorizes certainty into 4 levels:

- High: Further research is very unlikely to change our confidence in the estimate of effect.
- Moderate: Further research is likely to have an important impact on our confidence and may change the estimate.
- Low: Further research is very likely to have an important impact and is likely to change the estimate.
- Very Low: Any estimate of effect is very uncertain.

The assessment will consider risk of bias, consistency of results, directness of evidence, precision of estimates, and publication bias. We will create a Summary of Findings table presenting key information on included studies, synthesized results, and GRADE certainty assessments for each main outcome. The assessment will be done by two reviewers, with disagreements resolved by consensus or a third reviewer.

## Discussion

This systematic review will provide a comprehensive synthesis of the rapidly evolving literature on NLP methods for detecting delirium in hospitalized patients from clinical notes. By consolidating and critically appraising the available evidence, we aim to inform future research, development, and implementation efforts in this important area.

We will summarize the range of NLP architectures used, from rule-based algorithms to feature-engineered machine learning to deep learning approaches. Model performance will be compared across these categories, highlighting the potential advantages and limitations of each. We will examine differences in performance based on clinical setting, patient characteristics, note types, and labeling strategy.

We will identify the features and linguistic constructs that appear to be most informative for delirium detection across studies, potentially informing future feature engineering efforts. Common challenges in model development and evaluation will be discussed, along with innovative solutions proposed in the literature.

We will assess the completeness of reporting and risk of bias of individual studies, potentially identifying areas for improvement in the design and conduct of future work. We will also gauge the certainty of evidence for key outcomes using the GRADE framework, which will help contextualize the strength of conclusions that can be drawn from the review.

Based on the cumulative findings, we will outline key research gaps and propose high-priority future directions in NLP-based delirium detection. Considerations for successful clinical implementation and integration into real-world workflows will also be discussed.

This review will have several limitations. First, despite our comprehensive search strategy, we may miss some relevant studies, particularly in the grey literature or conference proceedings. However, we will attempt to mitigate this by contacting experts in the field and hand-searching reference lists of key articles.

Second, the anticipated heterogeneity in study designs, populations, settings, and NLP methods may preclude formal meta-analysis and limit the strength of conclusions that can be drawn. However, we will use a robust narrative synthesis approach to identify patterns, outliers, and moderators of model performance.

Third, our ability to assess risk of bias and completeness of reporting will be limited by the information provided in the primary studies. We will contact study authors for clarification where needed, but some uncertainties may remain. The addition of the TRIPOD checklist to our appraisal tools will help identify reporting gaps but will not eliminate this limitation entirely.

Finally, the rapid pace of progress in NLP and machine learning may result in the review findings becoming outdated quickly. To mitigate this, we will update the search prior to submission of the final review draft. We also see this review as a foundational step in an ongoing effort to synthesize and translate research in this dynamic field.

This systematic review will provide a rigorous, comprehensive evidence synthesis to guide the development and implementation of NLP-based delirium detection systems using clinical notes. The findings will identify the most promising technical approaches and highlight key challenges that require further research and innovation to overcome.

The results may inform the development of standardized benchmarks and reporting guidelines to enhance the quality and reproducibility of future NLP studies in delirium. The review may also uncover novel modeling strategies or data sources that have been underexplored to date.

From a clinical perspective, this review will lay the groundwork for the implementation and adoption of NLP-based delirium detection tools. By summarizing the performance of various approaches and identifying implementation considerations, the review may help health system leaders and policymakers make informed decisions about deploying NLP systems as part of multicomponent delirium programs.

Ultimately, this systematic review aims to advance the science and practice of delirium detection and management by harnessing the untapped potential of unstructured clinical data. By accelerating the development of accurate, generalizable, and implementable NLP models, this work may help improve the quality and safety of care for the many hospitalized patients at risk for delirium.

## Author Contributions

Ravi Shankar (Conceptualization, Data curation, Formal analysis, Investigation, Methodology, Project administration, Resources, Software, Supervision, Validation, Visualization, Writing—original draft, Writing—review & editing), Janani Kannan (Data curation, Formal analysis, Investigation, Methodology, Validation, Writing—review & editing), Yih Hng Tan (Data curation, Formal analysis, Investigation, Methodology, Validation, Writing—review & editing), and Xu Qian (Supervision, Writing—review & editing)

## Funding

This research received no specific grant from any funding agency in the public, commercial or not-for-profit sectors.

### Abbreviations

AUROC: area under the receiver operating characteristic curve
CAM: Confusion Assessment Method
CENTRAL: Cochrane Central Register of Controlled Trials
DSM-5: Diagnostic and Statistical Manual of Mental Disorders, Fifth Edition
EHR: electronic health record
GRADE: Grading of Recommendations, Assessment, Development and Evaluation
ICU: intensive care unit
NLP: natural language processing
NPV: negative predictive value
PPV: positive predictive value
PRISMA-P: Preferred Reporting Items for Systematic Review and Meta-Analysis Protocols
PROBAST: Prediction model Risk Of Bias ASsessment Tool
PROSPERO: International Prospective Register of Systematic Reviews

## Data Availability

Data sharing is not applicable to this protocol as no datasets were generated or analyzed. After completion of the review, the data extracted from the included articles will be made available in a public repository such as the Open Science Framework (https://osf.io/).

## Code Availability

Any custom code or software used to analyze data or generate results in the completed systematic review will be made available in a public repository such as GitHub (https://github.com/) to enable reproducibility and reuse.

## Appendix 1. Data Extraction Form

The following data items will be extracted from each included study using a standardized form in Covidence:

| Category | Variables |
| --- | --- |
| **Study Characteristics** | First Author, Year of Publication, Country, Funding Sources, Study Design |
| **Dataset** | Data Source(s), Time Period, Sample Size, Inclusion/Exclusion Criteria |
| **Patient Population** | Healthcare Setting, Mean Age (SD), Sex (% Female), Primary Diagnoses, Delirium Prevalence |
| **Clinical Notes** | Note Types Analyzed, Total Number of Notes, Average Number of Notes per Patient, Preprocessing Steps |
| **Delirium Definition** | Diagnostic Criteria, Screening Tools, Diagnostic Codes, Annotation/Labeling Process |
| **NLP Methods** | NLP Approach, Specific Algorithms, Software/Tools Used, Features Engineered, Vocabulary/Ontology Mapping, Handling of Negation and Uncertainty |
| **Model Performance** | Sensitivity, Specificity, Positive Predictive Value, Negative Predictive Value, F1 Score, AUROC |
| **Model Validation** | Internal Validation Methods, External Validation Cohorts, Sensitivity Analyses |
| **Implementation Considerations** | Computing Infrastructure, Training Time, Inference Time, Integration with EHR Systems, Interpretability/Explainability |
| **Key Conclusions** | Main Findings, Limitations, Implications for Practice and Research |

